# Impact of Ebola and COVID-19 pandemic on patterns in uptake of short-acting and long-acting contraceptive methods in Sierra Leone: a country-wide observational study

**DOI:** 10.1101/2025.02.04.25321692

**Authors:** Idrissa Mohamed Kamara, Bailah Molleh, Sulaiman Lakoh, Stephen Sevalie, Sartie M. Kenneh, Adrienne K. Chan, Sharmistha Mishra, Francis Moses, Abdulai Jawo Bah

## Abstract

**Background:** The Ebola epidemic and COVID-19 pandemic disrupted essential healthcare services globally, including family planning, but the extent of disruptions in Sierra Leone is largely unknown. This study aimed to evaluates the impact of these crises on the uptake of contraceptive methods—short-acting contraceptives (SAC) and long-acting reversible contraceptives (LARC)— among adolescent girls and women ages 10-45 years in Sierra Leone from 2013 to 2022.

**Methods:** We conducted a retrospective, cross-sectional study using aggregated DHIS2 data from adolescent girls and women aged 10-45 years. We describe and visualize the trends in the uptake of SAC and LARC before Ebola (March 2013 to April 2014), during Ebola (May 2014 to February 2016), after Ebola and before COVID-19 (March 2016 to February 2020), and during COVID-19 (March 2020 to February 2022). Proportional shifts and age-specific patterns were assessed to identify changes in contraceptive preferences.

**Results:** During the Ebola outbreak (2014–2016), SAC uptake declined temporarily while LARC uptake modestly increased. A recovery in SAC uptake followed during the inter-epidemic period (2016–2019). Conversely, the COVID-19 pandemic (2020–2022) saw an initial spike in SAC uptake, which subsequently returned to pre-pandemic levels. In contrast, LARC uptake consistently increased throughout both pandemics and the post-pandemic period. Notably, a proportional shift toward LARC methods was observed. Trends across age groups were variable, with adolescents demonstrating resilience in their use of contraceptives during the COVID-19 pandemic.

**Conclusion:** LARC services demonstrated remarkable stability during health emergencies, underscoring the resilience of long-term methods and the adaptability of women in making informed choices about their reproductive health amidst disruptions. These findings highlight the importance of strengthening healthcare systems to ensure continued access to reliable contraceptive options, particularly during crises.

## Introduction

Globally, 163 million women and adolescents face unmet needs for contraception; one-third of them are in sub-Saharan Africa (1). Fertility rates vary widely across sub-Saharan Africa (2). Countries in West Africa face some of the highest unmet needs (25%) with an estimated contraceptive prevalence of 15% (3). Modern contraceptive methods include short-acting contraceptive (SAC) methods (pills and injections) and long-acting reversible contraceptive (LARC) methods (intrauterine devices and implants) (4).

Sierra Leone, like many countries in sub-Saharan Africa, has a high maternal mortality rate of 443 per 100,000 live births ( 5 ). A key intervention to reduce maternal mortality is the prevention of unintended pregnancies. Currently, 25% of women of reproductive age in Sierra Leone do not have access to modern contraceptive services, and the proportion of demand met for modern contraception remains low (46%); with even fewer women in rural regions using modern contraception compared to urban women (6).

Gaps in contraceptive services in Sierra Leone may widen further during the public health emergency due to lockdown and movement restrictions. Sierra Leone was one of the countries severely affected by the 2014/2015 Ebola outbreak and subsequently suffered from the COVID-19 pandemic (7,8). During both of these public health emergencies, there were extensive interruptions in the delivery of health services including family planning services by interrupting the supply chain of contraceptives, diverting essential staff to support response efforts, and patient avoidance of hospital visits for fear of infections or lack of trust in the health system (9, 10).

The COVID-19 pandemic has highlighted the critical role of resilient family planning services, particularly the impact of contraceptive methods. In Senegal, women using short-acting contraceptives SACs faced greater challenges during lockdowns and supply chain disruptions compared to women using long-acting contraceptives (11). Unlike user-dependent methods such as SAC, which are prone to failure due to noncompliance, LARC does not rely on regular medication or visits to a clinic for injections (12, 13). Consequently, LARCs have demonstrated greater resilience in maintaining continuity of care during pandemic-related shocks and disruptions in health services (14).

Contraceptive use has been reported to be affected by the epidemic, but the extent to which long- and short-acting contraceptive methods have been affected has not been explored in Sierra Leone. This study aimed to describe patterns in the uptake of contraceptives and compare the uptake of LARC to uptake of SAC across a period spanning from the Ebola epidemic to the COVID-19 pandemic.

## Methods

### Study Design

We conducted a descriptive, retrospective, cross-sectional study using aggregated monthly national data from January 2013 to December 2022.

### Study Setting

Sierra Leone is located in West Africa. Its population is 7.5 million and has a fertility rate of 4.2 (7, 15). The country is divided into five geographic regions and 16 health districts. The health system has three levels-primary (community health centers, community health posts, and maternal and child health posts), secondary (district hospitals), and tertiary (specialized and referral hospitals).

Family planning services are provided in all facilities across the three tiers of the country’s healthcare system, free of cost to clients. Community Health Posts and Maternal and Child Health Posts mainly provide SAC and implant services, Community Health Centers provide SAC and LARC methods, and hospitals provide SAC, LARC, and permanent methods.

### Study Population

This study reviewed data from all adolescent girls and women aged 10-45 years who received contraception between March 2013 and December 2022. Sierra Leone experienced the Ebola epidemic between 2014 and 2016 and the COVID-19 pandemic between 2020 and 2022. We extracted data from March 2013 to March 2022 to assess the variation in the contraceptive uptake across the two public health emergencies over five time periods: March 2013 to April 2014 (pre-ebola), May 2014 to February 2016 (During Ebola), March 2016 to February 2020 (inter-epidemic-post-ebola and pre-COVID-19) and March 2020 to February 2022.

### Data Sources

On October 22, 2024, we extracted the monthly aggregated secondary data on family planning from DHIS2 from all 1,456 public and private health facilities providing reproductive health and family planning services across the country. Every month, data on the different family planning services at every facility were collated, summarised, and sent to the monitoring and evaluation officers at the district health management teams for validation and input into the DHIS2.

### Data Variables

We collected aggregate data on the number of newly enrolled contraceptives per month per 100 women aged 10-45 years, stratified by age group, district, and by method: LARC (intra-uterine device, implants, and SAC [injectable (intra-muscular or subcutaneous depo), oral contraceptive pills (combined oral contraceptives or progestin-only pills), emergency contraception, male condoms, and female condoms].

### Analyses

We used Stata (version 18) to conduct descriptive analyses of the monthly trends in SAC and LARC and the monthly median (interquartile range-(IQR) uptake of SAC and LARC in each time period. We compared the proportions of contraceptives that comprised SAC versus LARC and that of SAC and LARC by age group over the three time periods.

### Ethics consideration

Ethics approval was obtained from the Sierra Leone Ethics and Scientific Review Committee of the Ministry of Health (approval number 015/05/2023). Secondary data were abstracted under a waiver of informed consent, and the research team did not have access to information that could identify individual participants during or after data collection.

## Results

### Trends in the uptake of SAC and LARC among women aged 10-49 in Sierra Leone from 2013 to 2022

There were variations in the use of SAC between 2013 and 2016 (during the Ebola epidemic), with a slight increase in LARC use and a sharp decline in SAC use during the peak of the Ebola epidemic. Between 2016 and 2018, the use of SAC decreased, while the use of LARC gradually increased. There were fluctuations in the SAC uptake between 2018 and 2022, with a substantial increase in 2020-2021 and a decrease in 2021-2022. At the same time, LARC utilization continued to increase through the periods ( Figure 1 ).

**Figure 1:**
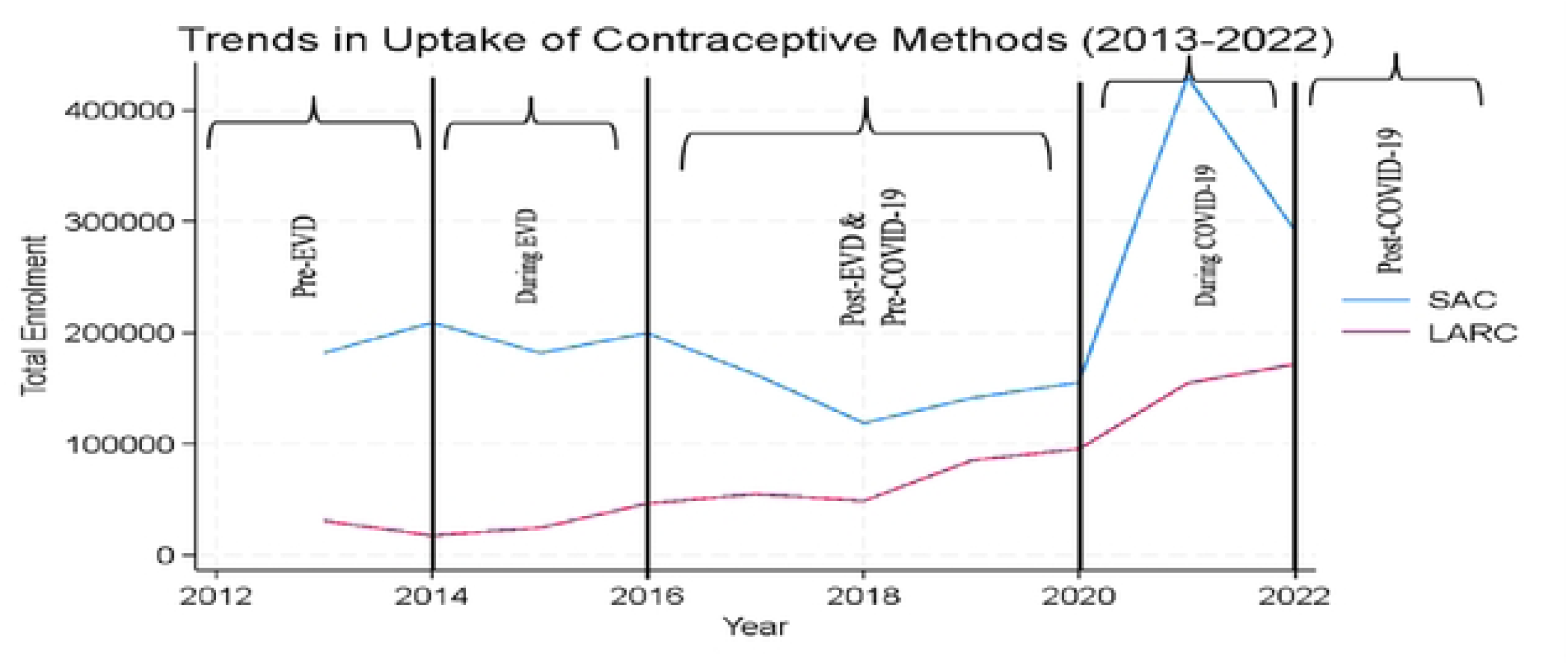
Trends in uptake of contraceptive methods (2013-2022) in Sierra Leone SAC=short acting contraceptives; LARC**=** long acting reversible contraceptives

### Trends in uptake of LARC and SAC pre, during, and after Ebola and pre, during, and after COVID-19

Before Ebola epidemic, the uptake of SAC was nine times higher than uptake LARC and remained at almost the same level during the outbreak. However, this proportion changed in favour of LARC uptake during the inter-epidemic period, remained constant throughout the COVID-19 epidemic period and drastically increasing during the post-COVID period (Figure 2).

**Figure 2:**
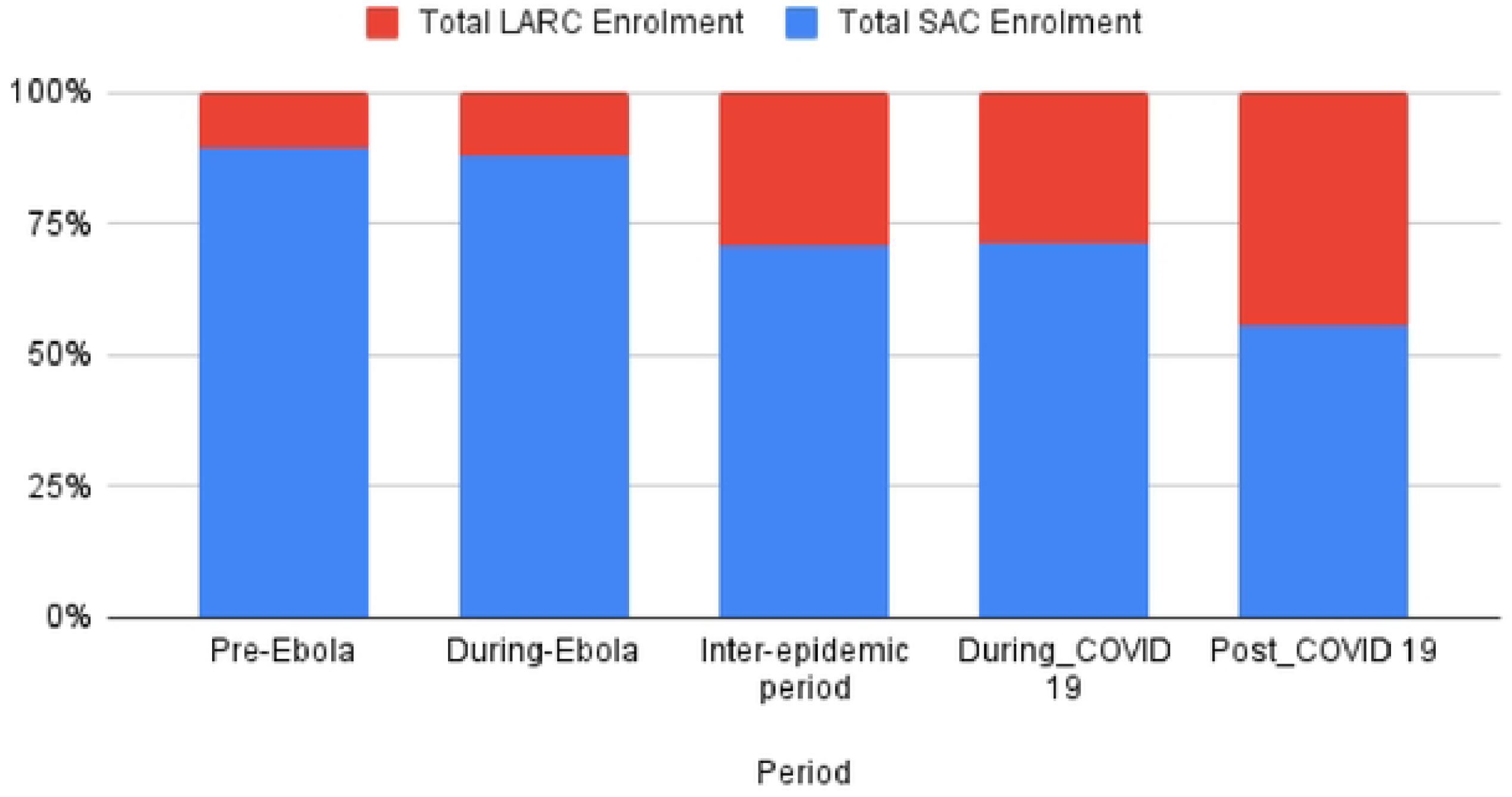
Proportion of SAC and LARC uptake during the different periods (2013-2022)

**Figure 3:**
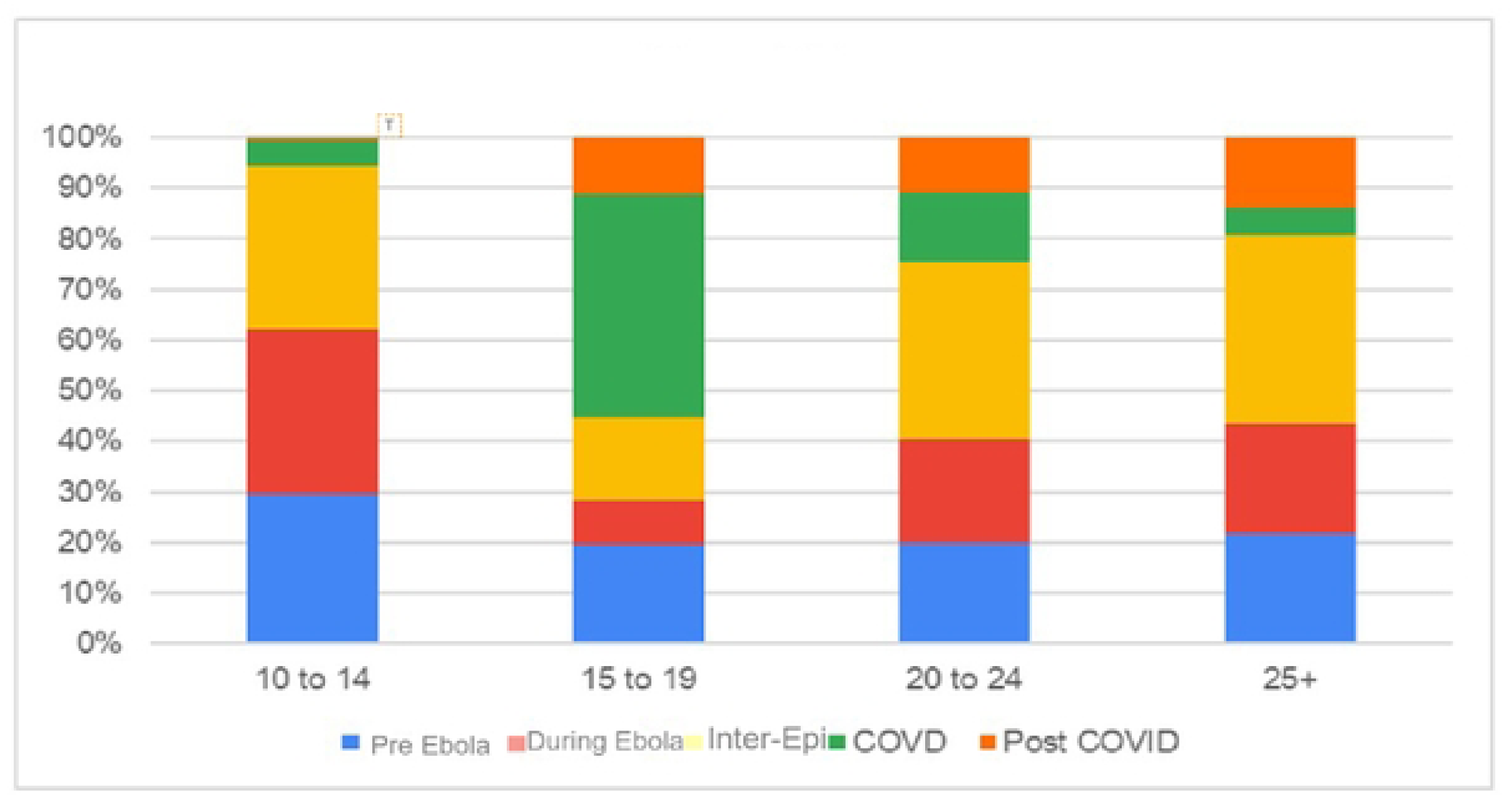
SAC uptake by different age groups at different period

**Figure 4:**
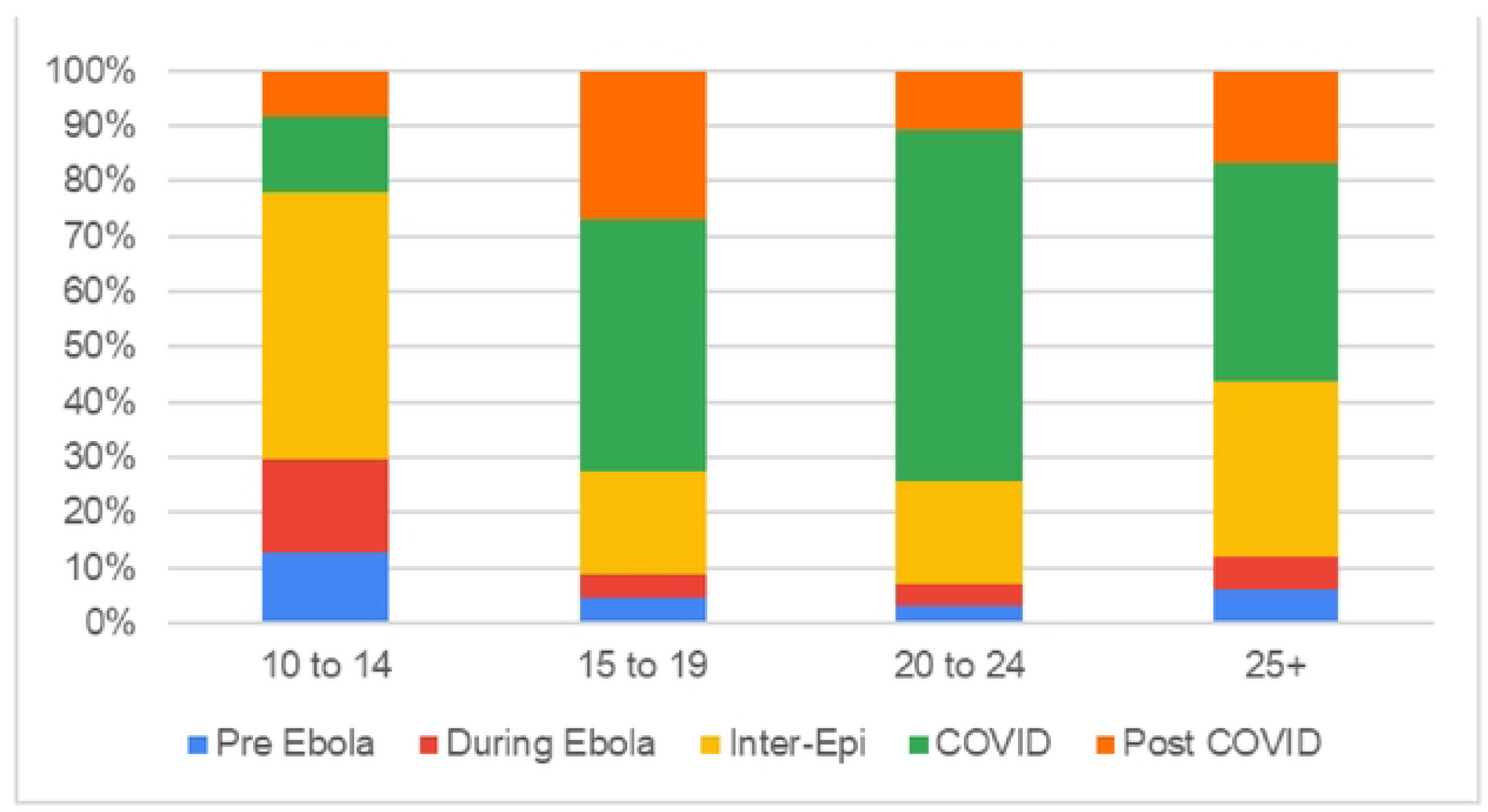
LARC uptake by different age groups at different periods

### The influence of age groups on the uptake of SAC and LARC pre-, during, and post-Ebola and pre-, during, and post-COVID-19

SAC uptake slightly increased in all age groups during the Ebola epidemic, except for those aged 15-19 years, where it decrease. Conversely, the COVID-19 pandemic saw a reduction in SAC uptake among all age groups, except for those aged 15-19 years, who exhibited a slight increase. In contrast, LARC uptake remained consistent across all age groups during the Ebola epidemic, showing resilience. However, at the onset of the COVID-19 epidemic, there was a shift toward an increase in LARC uptake among all age groups, except for people aged 25 years and above, where we observed a slight decrease in uptake.

## Discussion

In this first study on the impact of two public health emergencies on the use of contraceptive services in Sierra Leone between April 2013 and March 2022, we found that LARC uptake among women of reproductive age remained stable during the Ebola epidemic, whereas uptake increased in the post-Ebola epidemic and during the COVID-19 pandemic. Second, the 2014 Ebola epidemic and subsequent lockdowns and movement restrictions resulted in notable but brief interruption in the use of SACs. Third, there was a significant shift in preference from SAC to LARC uptake during the Ebola and COVID-19 periods. Finally, there were age variations in the uptake of SAC across the different time periods.

Our findings underscore the resilience of implementing LARC services in Sierra Leone amidst the challenges posed by the Ebola epidemic and the COVID-19 pandemic. Furthermore, the trends in the uptake of contraceptives (SAC and LARC) unveiled some insights into the impact of the Ebola epidemic and COVID-19 pandemic on the uptake of contraceptives in Sierra Leone, which may be informed by women’s adaptability in making informed choices about their reproductive health even in the face of unprecedented disruptions (16).

The sustained uptake of LARC during the Ebola and COVID-19 outbreaks reflects the improved access to and quality of family planning services ( 17 ). Data from the 2019 and 2008 Demographic Health Surveys showed an increasing trend in modern contraceptive use, from 6.7% in 2008 to 20.9% in 2019 (18). During the Ebola epidemic, there was a notable but brief interruption in the use of SACs, likely due to movement restrictions, widespread fear, and reallocation of resources from family planning and other basic healthcare needs (11, 19). However, at the start of the COVID-19 pandemic, SAC use surged threefold, but in 2021 it fell back to pre-pandemic levels. This corresponds with a spike in the uptake of LARC, which reduced slightly in 2021 but continued to increase in the post-COVID-19 period.

Our analysis highlight a considerable shift in preference from SAC to LARC uptake during the Ebola and COVID-19 period. LARC methods have a significantly higher uptake rate compared to SAC methods such as injectables and oral contraceptives. This shift may reflect evolving fertility preferences, programmatic investment in training and supply of LARC commodities and equipment, lesser side effects of LARC, and a growing preference for methods that require less frequent resupply and facility visits, or a shift towards more effective and long-term contraceptive options (20). In Burkina Faso and Kenya, the majority of women did not change their contraceptive methods during the initial stages of the pandemic, but among those who did make a switch, there was a tendency to opt for more effective long-acting contraceptive methods (21).

During the Ebola epidemic, we observed a slight increase in SAC uptake in all age groups, except for those aged 15-19 years, in which SAC uptake decreased. This finding suggests that despite the challenges posed by the Ebola outbreak, uptake of SAC remained high across most age groups, highlighting the importance of maintaining access to essential reproductive health services during emergencies (22). In contrast, the the COVID-19 pandemic led to a decrease in SAC uptake in all age group, except for the 15-19 age group, which saw a slight increase. This decline in SAC uptake during the COVID-19 pandemic underscores the disruptive impact of the pandemic on contraceptive services, particularly among older population. The observed increase in SAC uptake among adolescents may reflect unique challenges and needs faced by this age group during the pandemic, such as disruptions in education and limited access to traditional sources of support and information.

Finally, our study found that LARC uptake remained consistent across all age groups during the Ebola epidemic, demonstrating a remarkable resilience in the uptake of long-acting contraceptives (23). However, with the advent of the COVID-19 pandemic, there was a shift towards an increase in LARC uptake among all age groups, except for individuals aged 25 years and above, where a slight decrease in uptake was observed. This shift in LARC uptake patterns during the COVID-19 pandemic may indicate a greater preference for longer-acting contraceptive methods that require fewer clinic visits and offer longer-term protection against unintended pregnancies.

Our study has limitations. First, the dataset used for this study only contains information from public health facilities. The absence of data from additional sources such as pharmacies and private healthcare providers hinders the comprehensive assessment of contraceptive uptake. Second, the accuracy of data entry during lockdowns and movement restrictions may have affected the quality of data in the DHIS2, and thus the reliability of the data during those specific timeframes.

## Conclusion

Despite significant disruptions, LARC services demonstrated remarkable stability during health emergencies, underscoring the resilience of long-term methods and the adaptability of women in making informed choices about their reproductive health amidst disruptions. These findings highlight the importance of strengthening healthcare systems to ensure continued access to reliable contraceptive options, particularly during crises. Policy recommendations include expanding access points, diversifying contraceptive providers, and bolstering private sector involvement to improve family planning resilience in Sierra Leone. Furthermore, consideration of exploring urban-rural and inter-district differences in future studies can help guide government and implementing partners on the allocation of resources to support contraceptive services in public health emergencies.

## Declaration section

The authors declared no conflict of interest

## Acknowledgements

We thank Kristy Yiu (Unity Health Toronto) for her early support in coordination of project. SM is supported by a Tier 2 Canada Research Chair in Mathematical Modeling and Program Science.

## Author’s contribution

Conceptualization and study design: IMK, FM, AJB and SL

Data curation and validation: IMK, FM, SM, AKC

Methodology and formal analysis: IMK, SM, AJB, BM

Funding acquisition, resources, supervision: SS, SL, SM, AKC

Writing – original draft preparation: IMK, SMK, FM, AJB, AKC and SM

Writing – review & editing: SL, AKC and SM

## Availability of data

The datasets used and/or analyzed (10.6084/m9.figshare.28302206) for this study are available from: https://figshare.com/account/items/28302206/edit.

